# Blood pressure, cardiometabolic traits and cardiovascular events in women with uterine fibroids: a genetic correlation and Mendelian randomization study

**DOI:** 10.1101/2024.04.05.24305381

**Authors:** Joséphine Henry, Takiy Berrandou, Lizzy M. Brewster, Nabila Bouatia-Naji

## Abstract

**Background:** Uterine fibroids (UFs) are under-studied uterus neoplasms, affecting women of reproductive age and often leading to hysterectomy. Clinical series suggest impaired cardiometabolic features including hypertension in UFs. We investigated potential genetic links between blood pressure (BP), several cardiometabolic traits and events and UFs.

**Methods:** We used summary statistics of genome-wide association studies (GWAS) for UFs and 18 traits related to BP and cardio-metabolism. We applied linkage disequilibrium score regression to estimate genetic correlations and GCTA-mtCOJO for adjusted correlations. Univariate and bi-directional Mendelian randomization (MR) were used to test causal associations with UFs. We computed inverse variance-weighted. Weighted median estimation and MR-Egger regression were computed for sensitivity analyses. Multiple testing was addressed by Bonferroni correction.

**Results:** UFs significantly correlated with systolic (r_g_=0.08, *P*=8.7×10^−5^) and diastolic (r_g_=0.12, *P*=8.2×10^−8^) BP, including after adjustment on body mass index (BMI). UFs positively corelated with BMI (r_g_=0.11, *P*=4.1×10^−4^), waist-to-hip ratio (WHR) (r_g_=0.09, *P*=7.3×10^−3^), diabetes (r_g_=0.15, *P*=1.9×10^−5^) and triglycerides (TG) (r_g_=0.17, *P*=7.6×10^−7^). We identified a negative correlation with sex hormone-binding globulin (SHBG) (r_g_=-0.16, *P*=3×10^−4^), a marker of bio-availability of sex-steroids. We found no evidence for shared genetics with vascular diseases, except migraine (r_g_=0.08, *P*=5.8×10^−7^). MR analyses supported BMI, WHR, TG and SHBG, to causally associate with increased risk for UFs.

**Conclusions:** Our study shows that UFs share substantial genetic basis with traits related to BP, obesity, diabetes, in addition to migraine, a predominantly female vascular condition. We provide MR-based evidence for central obesity, visceral fat traits and sex-steroids bio-availability as relevant genetic risk factors for UFs.

## Introduction

Uterine fibroids (UFs), also known as uterine leiomyomata, are benign smooth muscle cell tumors of the uterus, affecting mainly women of reproductive age ^1^. Although they are mostly asymptomatic, UFs cause a large range of gynecological symptoms including abnormal uterine bleeding and issues with fertility, and less frequently, constipation and dyspareunia ^2^. The prevalence of UFs ranges from 4.5 to 68.6% depending on the studies ^3^. UFs accounts for up to 60% of hysterectomies, 30% among women under 44 years ^4^. Recently, techniques requiring less invasive surgery are becoming more popular in the treatment of UFs such as myomectomy and uterine arteries embolization ^2^.

Several factors were reported to influence the risk of developing UFs. Increasing parity is reported to be a protective factor for UFs, while early menarche and the use of oral contraceptives are suspected risk factors ^5^. Other risk factors include increasing pre-menopausal age, estrogen and testosterone levels, in addition to unhealthy diet and lifestyle ^6^. The precise mechanisms behind the association with ancestry, with relatively low occurrence in women of European ancestry, intermediate in Asian, and higher occurrence in women of African ancestry, are currently being explored ^7,8^.

Molecular and genetic explorations of UFs smooth muscle cell tumors indicate the role of driver mutations and common genetic variants in recurrent molecular pathways. The most prevalent somatic mutations were reported in the RNA Polymerase II (Pol II) mediator subunit gene (*MED12*) ^9,10^. In lower frequencies, mutations were also reported in the high mobility group AT-hook 2 (*HMGA2*) group, among others ^11^. To date, genome-wide association studies (GWAS) identified several common genetic variants to associate to the risk of UFs ^12^. Interestingly, some of the risk variants locate nearby *MED12* and *HMGA1*, belonging to the same gene family as *HMGA2* previously involved in fibroid growth via upregulation of proto-oncogenes ^13^.

A comprehensive investigation of the potential genetic links between UFs and its cardiometabolic risk factors is missing. UFs have been clinically associated with higher body mass index (BMI) and blood pressure ^8,14^, suggesting a potential role of cardiometabolic risk factors in their etiology. Nonetheless, the role of genetic determinants of major cardiovascular risk factors, including hypertension, has never been explored so far for the risk of UFs. Our study aims to explore the genetic links between a large panel of cardiometabolic traits and diseases and the risk for UFs through the application of genetic epidemiology methods. Through linkage disequilibrium score regression (LDSC) we estimated genetic correlations with UFs for blood pressure, lipids and metabolic traits and several major cardiovascular events. We also applied Mendelian Randomization (MR) based methods to examine the potential causal associations of correlated traits and diseases with UFs.

## Methods

### Access to publicly available GWAS data

The GWAS summary statistics used in this study were obtained from several publicly available resources (Supplementary Table 1). Data for UFs were obtained from Gallagher et al. ^12^, a large meta-analysis containing approximately 35,474 cases and 267,505 controls from four population-based studies and one direct-to-consumer cohort. Summary statistics for the 18 cardiometabolic traits were obtained from several published GWAS and the UK Biobank datasets available on (http://www.nealelab.is/uk-biobank). For each summary statistics, a quality control was applied and only variants fulfilling the following criteria were included: minor allele frequency (MAF) >0.01, info score (R^2^) > 0.9, Hardy-Weinberg equilibrium test p-value > 10^−6^, and biallelic variants.

**Table 1:**
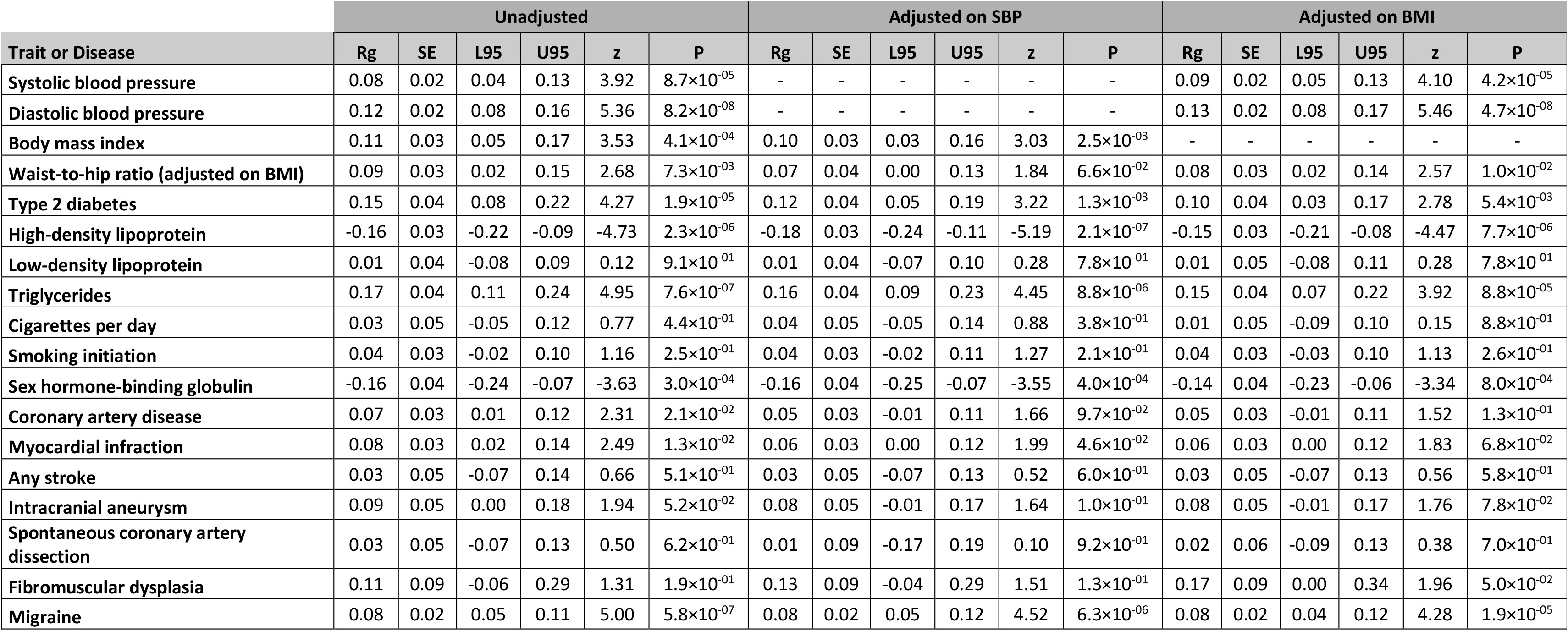
Genetic correlation between uterine fibroids and cardiometabolic traits and diseases. The table displays genetic correlations computed using LD score regression (Unadjusted) and the genetic correlation after GCTA mtCOJO adjustments on SBP and BMI. Rg: genetic correlation estimate, SE: standard error, L95: lower bound of the 95% confidence interval, U95: upper bound of the 95% confidence interval, z: z-score of, P: p-value.

### SNP-based genetic correlation analyses

We used the linkage disequilibrium score regression (LDSC) method (v1.0.1, [https://github.com/bulik/ldsc]) to assess genetic correlations with cardiometabolic traits and diseases, adhering to the standard algorithm recommended by the developers ^15,16^. Summary statistics for each trait were filtered to include only HapMap III SNPs panel, aiming to minimize bias from suboptimal imputation quality. Our analysis was limited to summary statistics from European ancestry GWAS, using files of linkage disequilibrium (LD) score in Europeans derived from the 1000 Genomes reference panel, as provided by the developers. To address potential sample overlap from the comprehensive UK Biobank dataset and prevent inflated false-positive rates, we did not constrain the LD score regression intercept in our estimates. Given the extensive range of traits examined, we applied the Bonferroni correction to account for multiple testing across 18 clinically independent traits, setting a strict significance threshold of *P* < 0.0027 (0.05/18). We also implemented the multi-trait-based conditional and joint analysis (mtCOJO) tool from the GCTA pipeline ^17^ to adjust GWAS summary statistics for uterine fibroids by factoring in systolic blood pressure (SBP) and BMI. These adjusted summary statistics were then employed to re-assess the genetic correlations between uterine fibroids, conditioned on SBP or BMI, and other traits of interest. This step served as a sensitivity analysis to evaluate the impact of SBP and BMI on the genetic correlation of uterine fibroids with these traits.

### Mendelian randomization analyses

We applied univariate and bi-directional Mendelian randomization (MR) settings as summarized in Supp figure 1 using MendelianRandomization (v0.6.0) and TwoSampleMR (v0.5.6) R (v4.0.4) packages, respectively. To ensure the robustness of our MR analyses, we implemented a meticulous selection process for instrumental variables (IVs), adhering to the three core MR assumptions: (1) a strong association with the exposure; (2) independence from confounders affecting both the exposure and outcome; and (3) an effect on the outcome solely through the exposure. This was achieved by employing LD clumping with stringent criteria (P-value threshold < 5 × 10^−8^, LD r^2^ < 0.001 within a 10,000 kb window) based on the European population data from the 1000 Genomes Project.

**Figure 1:**
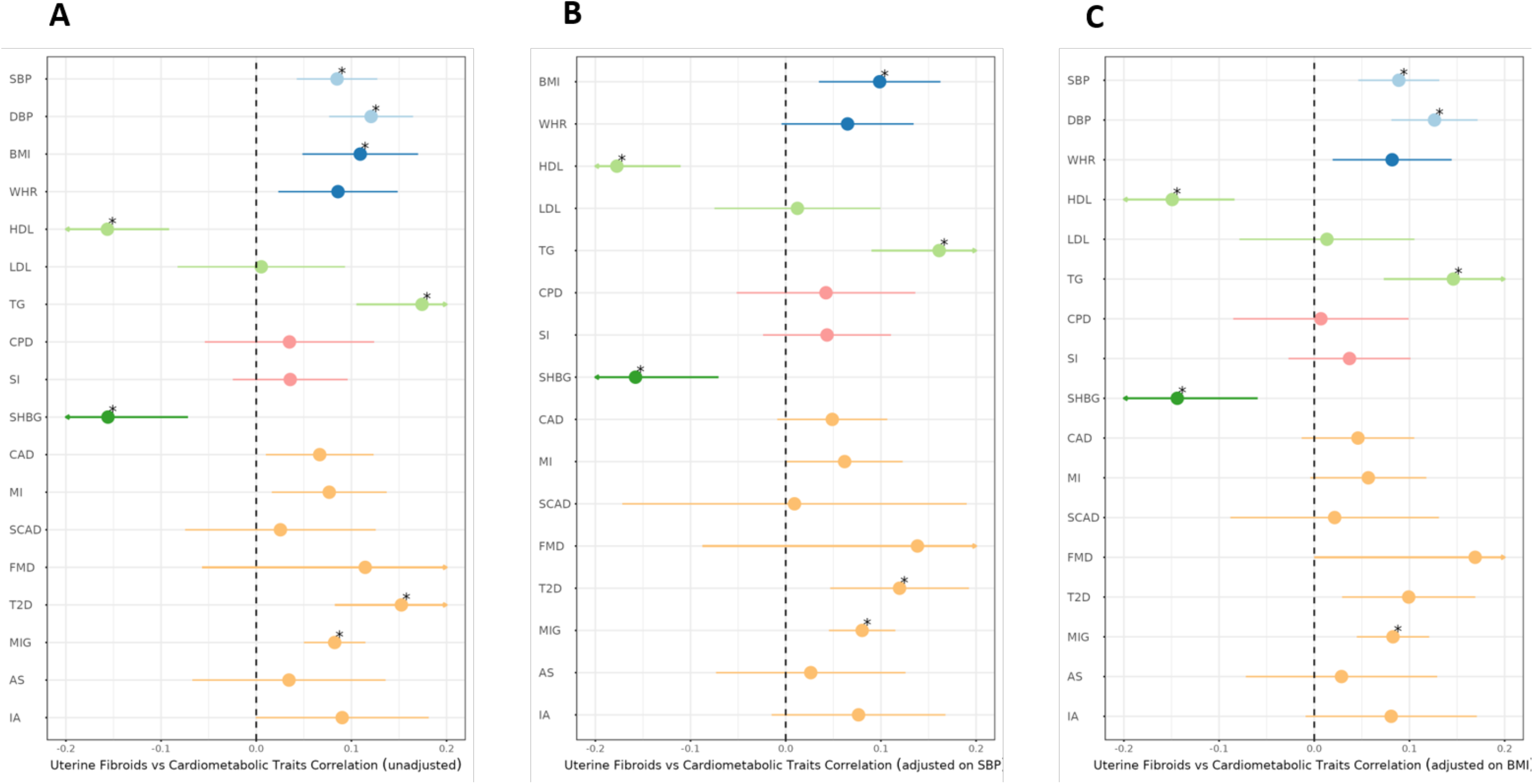
Forest plots of the genetic correlations between uterine fibroids and cardiometabolic traits. Rho coefficient of genetic correlation (Rg) is represented on the x-axis and range represents the 95 % CI. *P*-value of correlation for 18 tests adjusted using Bonferroni method is indicated as: * *P*.*adj* <0.05. **(A)** represents the genetic correlations between uterine fibroids and cardiometabolic traits unadjusted. **(B)** represents the genetic correlations between uterine fibroids and cardiometabolic traits adjusted on systolic blood pressure. **(C)** represents the genetic correlations between uterine fibroids and cardiometabolic traits adjusted on body mass index.

For the estimation of associations between genetically predicted risk factors and outcomes, we used the multiplicative random-effects inverse variance-weighted (IVW) method, considered as the main two-sample MR method, as recommended in the absence of evidence of pleiotropy in MR analyses ^18^, with estimates scaled to reflect clinically meaningful changes. Sensitivity analyses, including weighted median estimation and MR-Egger regression ^18,19^, were conducted to assess the consistency of our findings under different assumptions regarding genetic pleiotropy. These analyses are crucial for validating the MR results by providing evidence on the robustness of the IVW method’s estimates against potential pleiotropic effects. Furthermore, heterogeneity among the estimates was evaluated using Cochran’s Q test, ensuring the homogeneity of IV effects across different variants.

To correct for multiple testing, given our assessment of 9 risk factors, we applied a Bonferroni correction, setting a stringent significance threshold of *P* < 0.0055 for CVD traits and *P* < 0.006 for bidirectional MR involving 8 diseases. This adjustment reflects a conservative approach to significance, acknowledging the multiple comparisons made in our analysis. Results yielding P-values between 0.0055 and 0.05 were considered suggestively significant, indicating trends that may warrant further investigation.

## Results

### Uterine fibroids share a common genetic basis with blood pressure and several cardiometabolic traits

We first explored the genetic correlations between UFs and a set of traits related to blood pressure (BP), body weight and shape using LDSC ^15^ (Figure 1A, Table 1). We found UFs to correlate significantly with systolic (SBP, r_g_ = 0.08, *P* = 8.7×10^−5^) and diastolic (DBP, r_g_ = 0.12, *P* = 8.2×10^−8^) blood pressure, BMI (r_g_ = 0.11, *P* = 4.1×10^−4^) and waist-to-hip ratio (WHR) (r_g_ = 0.09, *P* = 7.3×10^−3^). SBP and DBP correlations remained unchanged after adjustment on the genetics of BMI using the GCTA mtCOJO method (r_gSBPadjBMI_ = 0.09, *P* = 4.2×10^−5^ and r_gDBPadjBMI_ = 0.13, *P* = 4.7×10^−8^ respectively, Figure 1B-C, Table 1). The genetic correlation between UFs and BMI also remained significant when we adjusted on SBP genetic results but did not survive multiple testing correction (r_gBMIadjSBP_ = 0.10, *P* = 2.5×10^−3^).

We then investigated the shared genetic basis between UFs and several metabolic diseases and traits. UFs and type 2 diabetes (T2D) seem to share a significant proportion of their genetic basis (r_g_ = 0.15, *P* = 1.9×10^−5^, Table 1). This correlation was only marginally affected by the adjustment on both SBP and BMI GWAS results (Table 1). A positive genetic correlation was established with triglycerides (TG) levels (r_g_ = 0.17, *P* = 7.6×10^−7^), a marker of metabolic dysregulation and liver fat accumulation, and a negative correlation with high-density lipoprotein (HDL) cholesterol (r_g_ = −0.16, *P* = 2.3×10^−6^) (Figure 1A, Table 1 and Supp table 2). These correlations remained unchanged after mtCOJO adjustments on both SBP and BMI GWAS results (Figure 1B-C, Table 1 and Supp table 2). A significant negative genetic correlation was detected with sex-hormone-binding globulin (SHBG) levels (r_g_ = −0.16, *P* = 3×10^−4^), a blood glycoprotein produced in the liver that regulates bioavailability of sex-steroids, including after adjustments on SBP and BMI genetics (Figure 1, Table 1 and Supp table 2).

**Table 2:** Mendelian randomisation (MR) analyses between UFs and cardiovascular risk factors, hormones and blood traits. N SNP: number of SNPs used as instrumental variables in the MR analysis, BETA: effect size obtained from the inverse variance weighted (ivw), Egger or weighted median methods, SE: standard error of effect size, P: p-value.

| Outcome : UF |  |  |  |  |  |  |  |  |  |  |  |  |  |
| --- | --- | --- | --- | --- | --- | --- | --- | --- | --- | --- | --- | --- | --- |
| Exposure (risk factor) | N SNP | BETA (IVW) | SE (IVW) | P (IVW) | BETA (Egger) | SE (Egger) | P (Egger) | BETA (weighted median) | SE (weighted median) | P (weighted median) | Intercept | Intercept SE | Intercept P value |
| Systolic blood pressure | 443 | 0.004 | 0.002 | $8.5 \times 10^{-02}$ | -0.002 | 0.007 | $7.3 \times 10^{-01}$ | 0.00 | 0.003 | 1.0 | 0.002 | 0.002 | $3.1 \times 10^{-01}$ |
| Diastolic blood pressure | 456 | 0.006 | 0.004 | $1.5 \times 10^{-01}$ | -0.01 | 0.010 | $1.8 \times 10^{-01}$ | 0.002 | 0.006 | $7.6 \times 10^{-01}$ | 0.004 | 0.002 | $4.0 \times 10^{-02}$ |
| Body mass index | 267 | 0.03 | 0.008 | $6.1 \times 10^{-05}$ | -0.01 | 0.023 | $5.6 \times 10^{-01}$ | 0.02 | 0.014 | $1.6 \times 10^{-01}$ | 0.005 | 0.002 | $3.5 \times 10^{-02}$ |
| Waist-to-hip ratio (adjusted on BMI) | 333 | 0.19 | 0.047 | $3.3 \times 10^{-05}$ | 0.29 | 0.108 | $6.6 \times 10^{-03}$ | 0.12 | 0.062 | $4.6 \times 10^{-02}$ | -0.002 | 0.002 | $3.0 \times 10^{-01}$ |
| High-density lipoproteins | 187 | -0.12 | 0.105 | $2.5 \times 10^{-01}$ | 0.12 | 0.176 | $4.8 \times 10^{-01}$ | 0.10 | 0.143 | $5.1 \times 10^{-01}$ | -0.003 | 0.002 | $8.6 \times 10^{-02}$ |
| Triglycerides | 147 | 0.16 | 0.038 | $1.9 \times 10^{-05}$ | 0.14 | 0.063 | $2.3 \times 10^{-02}$ | 0.20 | 0.055 | $4.0 \times 10^{-04}$ | 0.001 | 0.002 | $6.9 \times 10^{-01}$ |
| Cigarettes per day | 12 | 0.05 | 0.075 | $5.1 \times 10^{-01}$ | 0.08 | 0.167 | $6.2 \times 10^{-01}$ | -0.05 | 0.088 | $6.0 \times 10^{-01}$ | -0.002 | 0.008 | $8.1 \times 10^{-01}$ |
| Smoking initiation | 69 | -0.05 | 0.062 | $3.8 \times 10^{-01}$ | -0.12 | 0.304 | $6.8 \times 10^{-01}$ | -0.07 | 0.083 | $4.0 \times 10^{-01}$ | 0.002 | 0.008 | $8.1 \times 10^{-01}$ |
| Sex hormone-binding globulin | 162 | -0.005 | 0.002 | $2.5 \times 10^{-03}$ | -0.008 | 0.003 | $1.1 \times 10^{-02}$ | -0.01 | 0.002 | $2.7 \times 10^{-02}$ | 0.002 | 0.002 | $2.8 \times 10^{-01}$ |

Finally, we analyzed the genetic correlations between UFs and main forms of cardiovascular disease. We report a suggestively significant positive genetic correlation with coronary artery disease (CAD) (r_g_ = 0.07, *P* = 0.02). However, we found no support for a genetic link with stroke, intracranial aneurysm, or other vascular diseases with higher prevalence among women such as spontaneous coronary artery dissection (SCAD) ^20^ and fibromuscular dysplasia (FMD) ^21^ (Figure 1A, Table 1 and Supp table 2). The lack of significant genetic correlations was not masked by SBP and BMI genetic factors according to GCTA mtCOJO analyses (Figure 1B-C, Table 1 and Supp table 2). Interestingly, we report significant genetic correlation between UFs and migraine (r_g_=0.08, *P*=5.8×10^−7^) that was only marginally affected by adjustment on SBP and BMI GWAS results (r_gMIGadjSBP_ = 0.08, *P* = 6.3×10^−6^, r_gMIGadjBMI_ = 0.08, *P* = 1.9×10^-5^, Figure 1, Supp Table 2).

### Central obesity and visceral fat markers as genetic risk factors for uterine fibroids

We applied Mendelian randomization (MR) to estimate the potential causal links between traits for which we reported significant genetic correlations and UFs. Two-sample MR analyses showed that genetically-predicted higher BMI (beta_IVW-BMI_=0.033±0.008, P=6.1×10^−5^) and WHR adjusted BMI (beta_IVW-WHR_=0.193±0.047, P=3.3×10^−5^), but not blood pressure traits, were associated with increased risk of UFs (Figure 2 and Table 2). Genetically-predicted higher TG levels were also associated with an increased risk for UFs (beta_IVW-TG_=0.163±0.038, P=1.9×10^−5^). Interestingly, genetically-predicted higher SHBG levels were associated with lower risk of UFs (beta_IVW-SHBG_=-0.005±0.002, P=2.5×10^−3^). However, bi-directional MR analyses indicated no evidence of reverse causality of genetic links with none of the diseases with which UFs shared their genetic basis (Supp figure 2 and Supp table 3).

**Figure 2:**
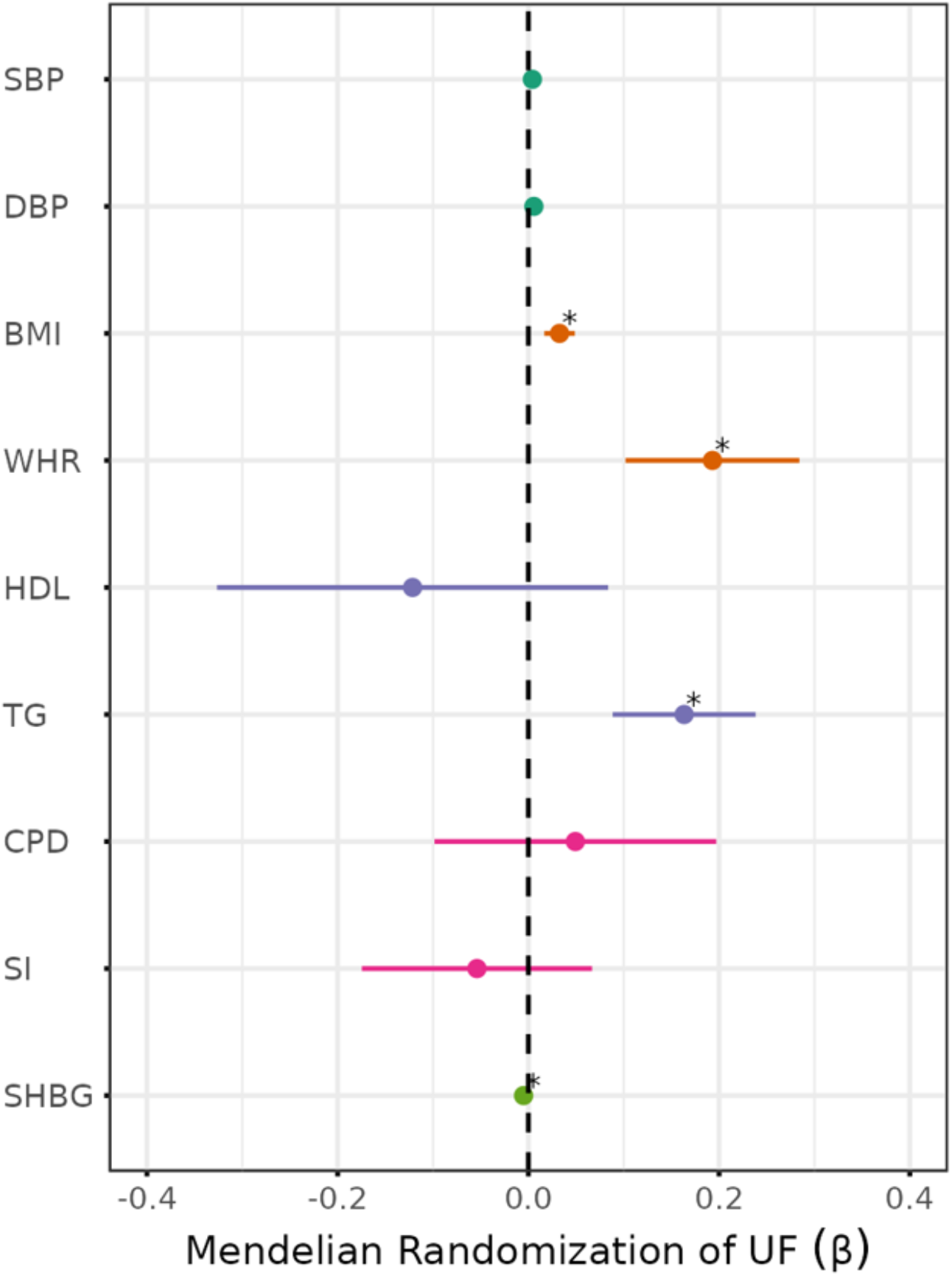
Forest plot of the Mendelian Randomization between uterine fibroids and its potential risk factors. Beta coefficient of Mendelian Randomization (−) is represented on the x-axis and range represents the 95 % CI. *P*-value of correlation for 9 tests adjusted using Bonferroni method is indicated as: * *P*.*adj* <0.05.

## Discussion

Our study shows that UFs share a substantial part of their genetic background with cardiometabolic traits related to blood pressure, obesity and diabetes in addition to migraine. Our Mendelian randomization findings do not support a causal relation between genetic predisposition to increased blood pressure. However, we report evidence for traits related to central obesity, including waist-to hip ratio, triglycerides and sex hormone binding globulin levels. We report these as relevant genetic risk factors for UFs supporting the role of visceral fat accumulation and its adverse metabolic consequences in UFs, an under-searched women specific condition affecting reproductive organs outside the context of pregnancy.

The present study reports a positive genetic correlation between UFs and both SBP and DBP. This result is in line with clinical reports highlighting that women with hypertension are at higher risk for UFs (7,8,21). The epidemiological co-occurrence of UFs with hypertension, and the positive genetic correlations we observed between blood pressure and UFs may mirror the existence of shared genetically determined mechanisms. Vascular cells proliferation and tissue remodeling share many biological mechanisms in the vascular wall and the uterine tissue. Both vascular and uterine smooth muscle cells can be induced by growth factors and vasoactive peptides ^22,23^, such as angiotensin-II, which was previously reported to induce the proliferation of UF cells *in vitro* ^24^. We have recently described ultrastructure abnormalities indicating degenerative changes in the contractile apparatus of smooth muscle cells, in resistance vessels of UFs patients, similar to fibroids abnormalities ^25^. However, our results do not support the existence of causal associations between genetically predicted fluctuations in blood pressure with the risk for UFs. The biological pathways linking blood pressure and UFs are complex and involve multiple intermediary factors or conditional metabolic and hormonal dependencies that were probably not captured by the current MR framework. Further population and molecular investigations are needed to disentangle the complex relation that may connect blood pressure to UFs.

Additionally, our study provides evidence for associations between genetically-determined central obesity and visceral fat content, and increased risk of UFs, supporting data from observational studies^22^.

Our data is confirmatory to recent findings where BMI and WHR were identified as important genetic risk factors for several female reproductive conditions, including UFs ^26^. Our data also provide first evidence of the role of genetically determined TG, a marker of visceral fat, in UFs risk in accordance with epidemiological evidence in the NFBC66 study, where one millimolar increase of circulating TG levels associated with 1.27 increase in the risk of UFs ^27^. Consistently, we found that genetically lowered SHBG levels are causally associated with a higher risk of development of UFs. The increase in visceral fat, especially around the liver where it is more biologically active ^28,29^, is a potential additional source of estrogen production. A genetically-determined lower availability or synthesis of SHBG by adipose tissue to counter-part the higher estrogen levels is a potential mechanism of uterine SMCs proliferation that may deserve deeper investigation ^22,29,30^.

Our results also show evidence for significant genetic correlation between UFs and T2D. Observational clinical data previously described higher risk of T2D among UF patients ^14,31^. However, the absence of causal association established by MR suggests that the two diseases share genetically determined biological mechanisms but the currently established genetic determinants of T2D has little influence on the genetic risk for UFs. One major suspected shared mechanism is central obesity, higher visceral fat content, altered lipid metabolism specifically in the liver and diminished bioavailability of SHBG. ^32–34^. Whether female sex-specific genetic risk factors for T2D may influence UFs onset may clarify this interesting genetic link between T2D and UFs.

Beyond blood pressure, our study does not support genetically driven causal relationship between cardiovascular diseases (CVDs) and UFs, despite evidence for UFs patients to be at higher cardiovascular risk ^8,14^. We report only nominally significant genetic correlations with coronary artery disease and myocardial infraction, suggesting a potential common genetic background with atherosclerotic disease. Suspected biological mechanisms are notably hyperlipidemia and SMCs remodeling, two major mechanisms of atherosclerotic disease ^31^. Nonetheless, we excluded any evidence of genetic correlations between UFs and fibromuscular dysplasia (FMD), a female arteriopathy, where SMCs proliferation is a suspected mechanism as reported in recent genetic investigation from our group (20). A potential explanation is the major discrepancy between UFs and FMD, with this latter not being influenced by dyslipidemia and overall or central obesity ^36^, whereas obesity is a known risk factor for UFs.

We report for the first time a genetic correlation between UFs and migraine. Recently, a significant genetic correlations was reported between endometriosis and migraine ^37^. These findings linking genetically migraine to several women’s reproductive diseases stresses further the need to dig into these shared molecular mechanisms, especially that migraine is a neurovascular disorder predominantly affecting women. The absence of association through MR between migraine and UFs suggests joint occurrence rather than causal relation. One possible explanation could be shared exposure to blood pressure and female-specific factors, such as hormones fluctuation during -the life course ^38^. Of note, ultrastructural phenotypic resemblance in vascular remodeling with UFs involving smooth muscle cells were described in superficial temporal and occipital arteries of patients with migraine ^25,39^.

The present study also presents several limitations. First, the UFs GWAS meta-analysis employed included overlapping samples from the UK Biobank that we used in some of the cardiometabolic datasets in the genetic correlation analyses, although we considered this situation in genetic correlation analyses through non-constraint on intercept. The lack of association causality through MR only reflects the variance of the traits controlled by genetic variants and not the overall variance of the traits analyzed. This study used only publicly available summary statistics, which limits the possibility to stratify on important potential confounding or mediating factors such as oral contraception intake, or parity. Our study was limited to genetic data generated mainly in women of European ancestry and may not be applicable in African or Asian ancestry, where UFs have higher prevalence. Overall, our findings need to be confirmed using larger and more diverse sets of patient cohorts.

In conclusion, our study confirms existing and delineates novel genetic links between the risk of UFs development and traits and diseases associated with cardiovascular risk. We demonstrate that UFs exhibit a substantial genetic overlap with blood pressure, central obesity, and T2D. Furthermore, we present evidence supporting mediation by shared mechanisms, notably smooth muscle cell remodeling, dyslipidemia, and impaired levels of sex hormone-binding globulin. Notably, we report, for the first time, a genetic correlation between UFs and migraine, likely attributable to shared yet not entirely overlapping genetic mechanisms involved in vascular remodeling with high relevance of blood pressure regulation, and potentially shared risk factors related to female sex. These findings unveil multiple avenues for further investigation to elucidate the etiology of UFs, an under-investigated and illy-understood women condition associated to poor cardiovascular health.

## Data Availability

All data produced in the present work are contained in the manuscript

## Sources of funding

This study was funded by the French Society of Cardiology, through Fondation Coeur et Recherche (to N.B.-N.); La Fédération Française de Cardiologie (to N.B.-N.).

## Disclosures

None

## Figures

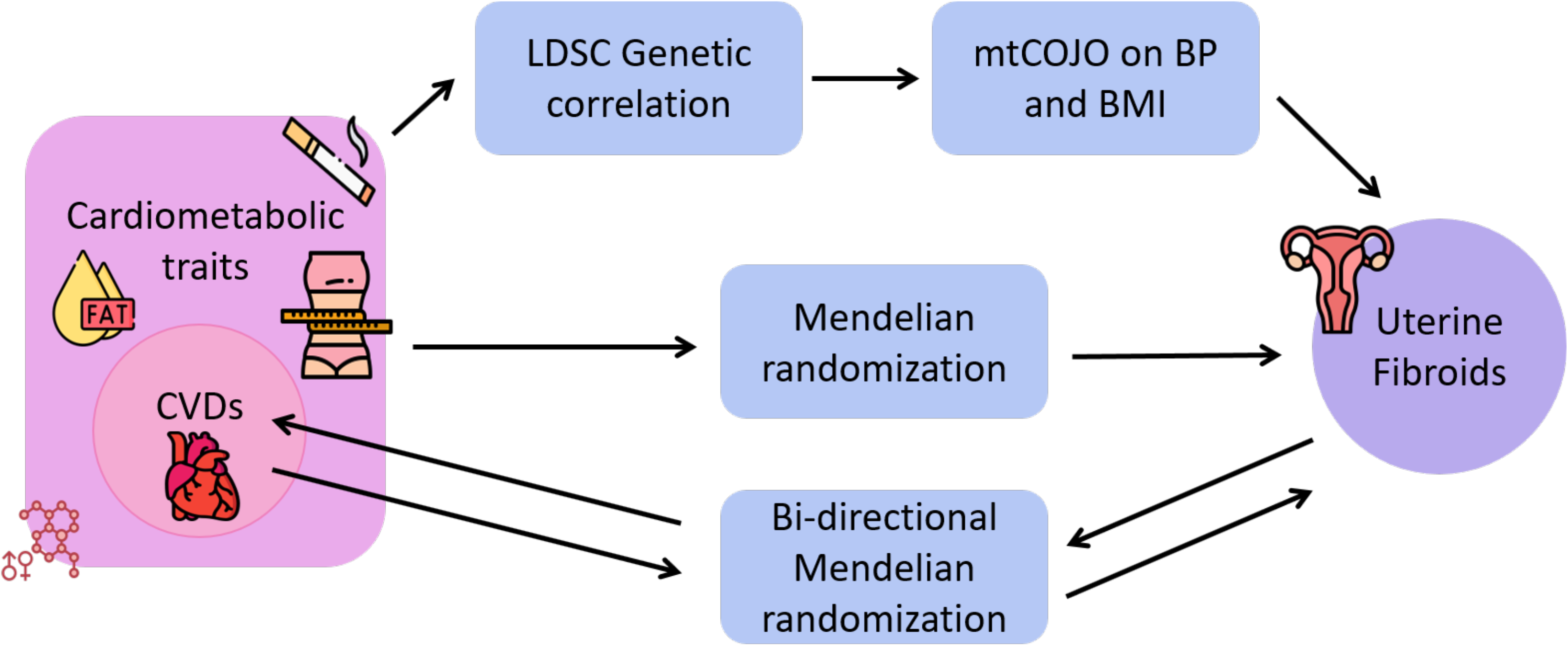

**Figure**: Study Abstract

## Supplemental Material

Tables S1–S2

Figure S1–S2

## Notes

### Competing Interest Statement

The authors have declared no competing interest.

### Funding Statement

This study was funded by the French Society of Cardiology, through Fondation Coeur et Recherche (to N.B.-N.); La Federation Francaise de Cardiologie (to N.B.-N.)

### Author Declarations

The study used (or will use) ONLY openly available human data that were originally located at : http://www.nealelab.is/uk-biobank https://broad-ukb-sumstats-us-east-1.s3.amazonaws.com/round2/additive-tsvs/21001_raw.gwas.imputed_v3.both_sexes.tsv.bgz https://broad-ukb-sumstats-us-east-1.s3.amazonaws.com/round2/additive-tsvs/30760_raw.gwas.imputed_v3.both_sexes.varorder.tsv.bgz https://broad-ukb-sumstats-us-east-1.s3.amazonaws.com/round2/additive-tsvs/30780_raw.gwas.imputed_v3.both_sexes.varorder.tsv.bgz https://broad-ukb-sumstats-us-east-1.s3.amazonaws.com/round2/additive-tsvs/30830_raw.gwas.imputed_v3.both_sexes.varorder.tsv.bgz http://ldsc.broadinstitute.org/ldhub/ https://genome.psych.umn.edu/index.php/GSCAN http://www.cerebrovascularportal.org

