## Supplemental tables and figures for "Blood pressure, cardiometabolic traits and cardiovascular events in women with uterine fibroids: a genetic correlation and Mendelian randomization study"

Henry et al.,

#### Supplementary tables

| Phenotype | Trait | N (Cohort or Cases / Controls) | Study reference (PMID) |
|---|---|---|---|
| Systolic blood pressure | SBP | [637578 – 745820] | 30224653 |
| Diastolic blood pressure | DBP | [647307 – 757601] | 30224653 |
| Body mass index | BMI | 359983 | UKBB (https://broad-ukb-sumstats-us-east-1.s3.amazonaws.com/round2/additive-tsvs/21001_raw.gwas.imputed_v3.both_sexes.tsv.bgz) |
| Waist-to-hip ratio (adjusted on BMI) | WHR | 697734 | 30239722 |
| High-density lipoprotein | HDL | 315133 | UKBB (https://broad-ukb-sumstats-us-east-1.s3.amazonaws.com/round2/additive-tsvs/30760_raw.gwas.imputed_v3.both_sexes.varorder.tsv.bgz) |
| Low-density lipoprotein | LDL | 343621 | UKBB (https://broad-ukb-sumstats-us-east-1.s3.amazonaws.com/round2/additive-tsvs/30780_raw.gwas.imputed_v3.both_sexes.varorder.tsv.bgz) |
| Triglycerides | TG | 343992 | UKBB (https://broad-ukb-sumstats-us-east-1.s3.amazonaws.com/round2/additive-tsvs/30870_raw.gwas.imputed_v3.both_sexes.varorder.tsv.bgz) |
| Cigarettes per day | CPD | 337334 | 30643251 |
| Smoking initiation | SI | 1232091 | 30643251 |
| Sex hormone-binding globulin | SHBG | 312215 | UKBB (https://broad-ukb-sumstats-us-east-1.s3.amazonaws.com/round2/additive-tsvs/30830_raw.gwas.imputed_v3.both_sexes.varorder.tsv.bgz) |
| Coronary artery disease | CAD | 60801 / 123504 | Cardiogram |
| Myocardial infarction | MI | 42561 / 123504 | Cardiogram |
| Stroke | AS | 40585 / 406111 | 29531354 |
| Intracranial aneurysm | IA | 7495 / 71934 | 33199917 |
| Migraine | MIG | 30465 / 143147 | 36939796 |
| Type 2 diabetes | T2D | 74125 / 824006 | 30297969 |
| Spontaneous coronary artery dissection | SCAD | 1917 / 9292 | 37248441 |
| Fibromuscular dysplasia | FMD | [1111 - 1578] / [5049 - 7100] | 34654805 |
| Uterine Fibroids | UF | 35474/ [190825 – 208850] | 31649266 |

**Supp. Table 1:** List of summary statistics leveraged in the study.

| Exposure | Outcome | N SNP | BETA (ivw) | SE (ivw) | P (ivw) | BETA (egger) | SE (egger) | P (egger) | BETA (weighted median) | SE (weighted median) | P (weighted median) | Directionality | Directionality P value |
|---|---|---|---|---|---|---|---|---|---|---|---|---|---|
| **Migraine** | **UFs** | 23 | -0,002 | 0,001 | $2,7\times10^{-01}$ | 0,004 | 0,003 | $2,1\times10^{-01}$ | -0,0004 | 0,002 | $8,3\times10^{-01}$ | TRUE | $1,4\times10^{-39}$ |
| **Coronary artery disease** | | 18 | -0,05 | 0,03 | $1,1\times10^{-01}$ | -0,006 | 0,10 | $9,6\times10^{-01}$ | -0,02 | 0,04 | $6,2\times10^{-01}$ | TRUE | $7,3\times10^{-60}$ |
| **Myocardial infraction** | | 18 | -0,03 | 0,03 | $4,7\times10^{-01}$ | 0,07 | 0,11 | $5,7\times10^{-01}$ | -0,01 | 0,04 | $7,6\times10^{-01}$ | TRUE | $8,5\times10^{-54}$ |
| **Type 2 diabetes** | | 23 | -0,02 | 0,03 | $6,0\times10^{-01}$ | -0,06 | 0,07 | $4,5\times10^{-01}$ | -0,04 | 0,03 | $1,8\times10^{-01}$ | TRUE | $1,2\times10^{-132}$ |
| **Any stroke** | | 23 | 0,03 | 0,03 | $3,4\times10^{-01}$ | -0,004 | 0,06 | $9,5\times10^{-01}$ | 0,02 | 0,04 | $6,7\times10^{-01}$ | TRUE | $3,5\times10^{-120}$ |
| **Intracranial aneurysm** | | 15 | -0,16 | 0,08 | $4,8\times10^{-02}$ | -0,003 | 0,26 | $9,9\times10^{-01}$ | -0,10 | 0,10 | $3,4\times10^{-01}$ | TRUE | $8,7\times10^{-04}$ |
| **Spontaneous coronary artery dissection** | | 11 | -0,11 | 0,24 | $6,6\times10^{-01}$ | -0,17 | 0,81 | $8,4\times10^{-01}$ | 0,07 | 0,26 | $8,0\times10^{-01}$ | FALSE | $8,7\times10^{-01}$ |
| **Fibromuscular dysplasia** | | 11 | 0,25 | 0,19 | $1,9\times10^{-01}$ | -0,04 | 0,59 | $9,4\times10^{-01}$ | 0,18 | 0,26 | $4,8\times10^{-01}$ | TRUE | $6,8\times10^{-01}$ |
| **UFs** | **Migraine** | 4 | -0,15 | 1,929 | $9,4\times10^{-01}$ | -6,98 | 7,08 | $4,3\times10^{-01}$ | 0,26 | 2,2 | $9,1\times10^{-01}$ | FALSE | $8,3\times10^{-01}$ |
| | **Coronary artery disease** | 22 | -0,01 | 0,04 | $7,8\times10^{-01}$ | -0,05 | 0,10 | $5,9\times10^{-01}$ | -0,03 | 0,05 | $4,5\times10^{-01}$ | TRUE | $7,7\times10^{-144}$ |
| | **Myocardial infraction** | 11 | -0,02 | 0,06 | $7,2\times10^{-01}$ | -0,06 | 0,13 | $6,6\times10^{-01}$ | -0,03 | 0,06 | $5,6\times10^{-01}$ | TRUE | $4,7\times10^{-121}$ |
| | **Type 2 diabetes** | 159 | 0,05 | 0,02 | $1,0\times10^{-02}$ | -0,01 | 0,04 | $7,5\times10^{-01}$ | 0,01 | 0,03 | $6,4\times10^{-01}$ | TRUE | $2,8\times10^{-02}$ |
| | **Any stroke** | 8 | -0,13 | 0,13 | $3,1\times10^{-01}$ | 0,69 | 0,93 | $4,9\times10^{-01}$ | -0,10 | 0,10 | $3,3\times10^{-01}$ | TRUE | $1,3\times10^{-41}$ |
| | **Intracranial aneurysm** | 7 | -0,03 | 0,03 | $3,6\times10^{-01}$ | -0,23 | 0,17 | $2,2\times10^{-01}$ | -0,01 | 0,04 | $7,6\times10^{-01}$ | TRUE | $1,7\times10^{-70}$ |
| | **Spontaneous coronary artery dissection** | 11 | 0,01 | 0,01 | $3,5\times10^{-01}$ | 0,12 | 0,03 | $5,7\times10^{-03}$ | 0,02 | 0,02 | $2,7\times10^{-01}$ | TRUE | $2,0\times10^{-102}$ |
| | **Fibromuscular dysplasia** | 1 | | | | | | | | | | | |

**Supp. Table 2:** Bi-directional Mendelian randomization (MR) analyses between UFs vs migraine and cardiovascular diseases.
N SNP: count of SNP used as instrumental variables in the MR analyses, BETA: effect size obtained from inverse variance weighted (IVW), Egger and weighted median MR analyses, SE: standard error of effect size, P: P-value for the association.

### Supplementary figures

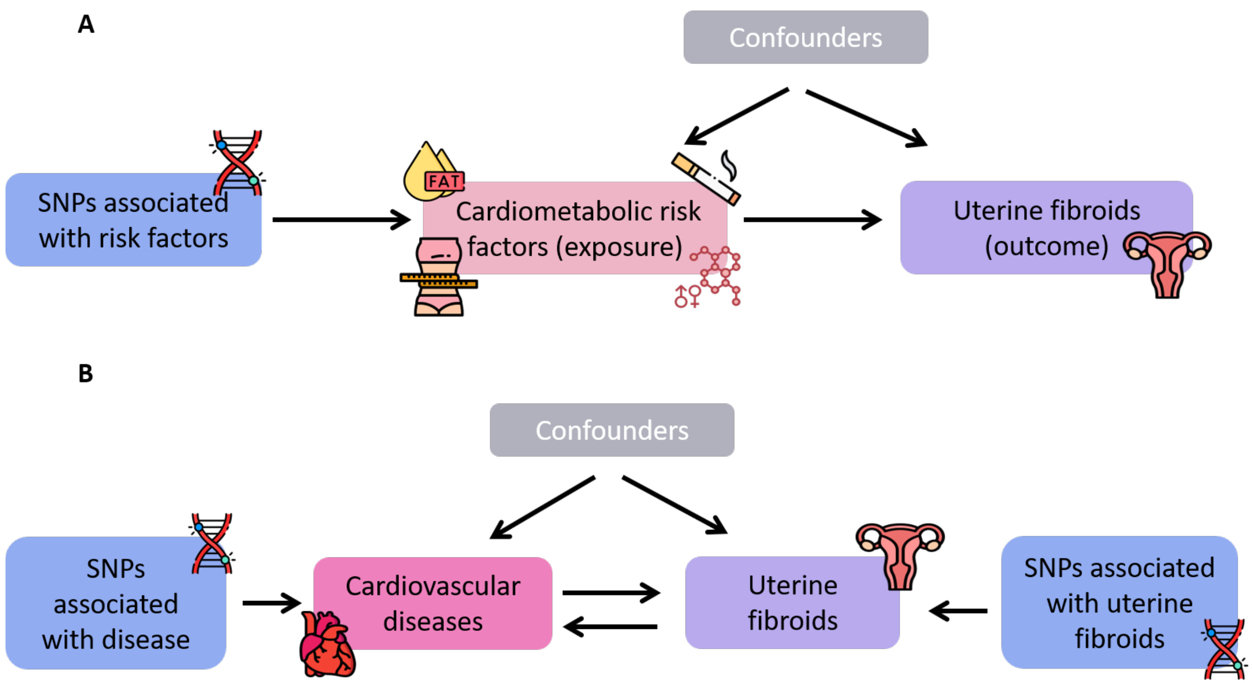


**Figure supp 1**: Schematic diagram of the different settings of Mendelian Randomization. **(A)** represents the univariate Mendelian Randomization analysis, used to estimate the causal effect of potential cardiometabolic risk factors. **(B)** represents the bi-directional Mendelian randomization analysis, used to estimate the reverse causality between uterine fibroids and cardiovascular diseases. The genetic variants used were selected with the GWAS significant threshold of $P<5\times10^{-8}$.

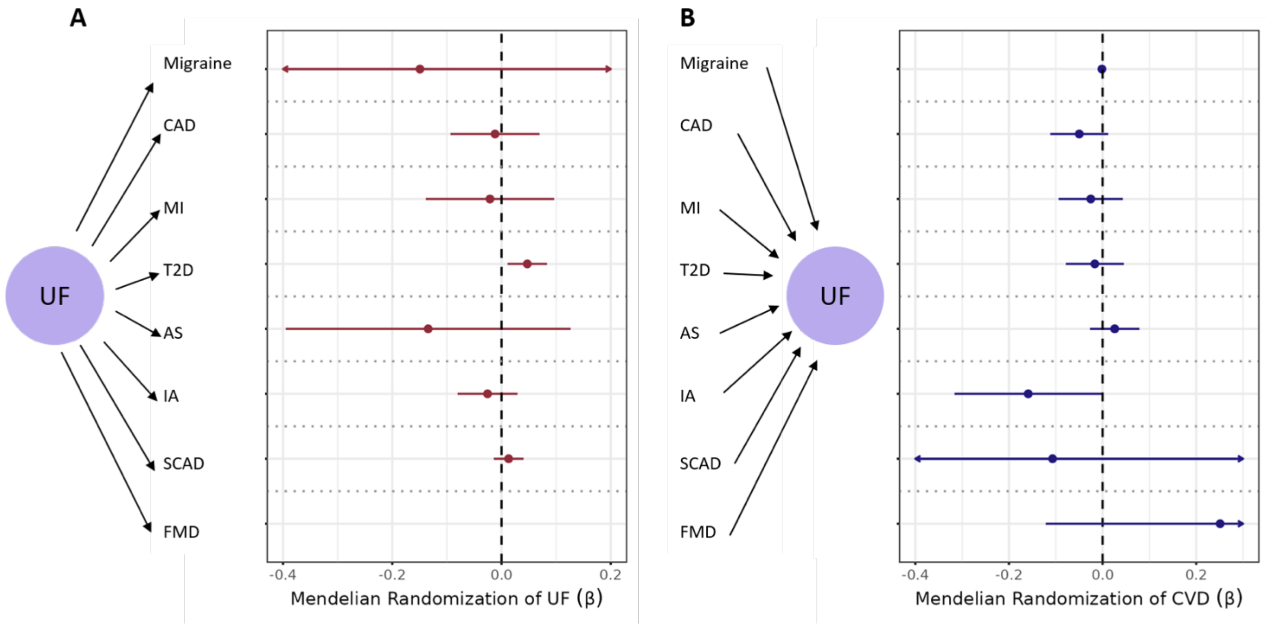


**Figure supp 2**: Forest plot of the bi-directional Mendelian Randomization between uterine fibroids and some cardiovascular diseases. Beta coefficient of Mendelian Randomization ($\boldsymbol{\beta}$) is represented on the x-axis and range represents the 95 % CI. *P*-value of association for 8 tests adjusted using Bonferroni method is indicated as: * *P.adj* <0.05. **(A)** represents the MR from UF to cardiovascular diseases. **(B)** represents the MR from cardiovascular diseases to UF.
